# Longitudinal tracking of intra-breath respiratory impedance in preschool children

**DOI:** 10.1101/2023.11.23.23298972

**Authors:** Matthew D Wong, Tamara Blake, Syeda Farah Zahir, Sadasivam Suresh, Zoltán Hantos, Keith Grimwood, Stephen B Lambert, Robert S Ware, Peter D Sly

## Abstract

**Background:** Longitudinal measurements of intra-breath respiratory impedance (Zrs) in preschool-aged children may be able to distinguish abnormal lung function trajectories in children with a history of wheezing compared to healthy ones.

**Methods:** Children from a prospective, longitudinal community-based cohort performed annual intra-breath oscillometry (IB-OSC) measurements from age 3-years to 7-years. IB-OSC was performed using a single 10 Hz sinusoid while clinically asymptomatic. Linear mixed-effects models were developed to explore the effects of wheezing phenotypes, growth, and sex on seven IB-OSC outcome variables over time: resistance at end-expiration (ReE), resistance at end-inspiration (ReI), the tidal change in resistance (ΔR=ReE-ReI), reactance at end-expiration (XeE), reactance at end-inspiration (XeI), the tidal change in reactance (ΔX=XeE-XeI), and ΔX normalised by tidal volume (ΔX/V_T_).

**Results:** Eighty-five children produced 375 acceptable IB-OSC measurements. Subjects were classified into one of three wheeze groups: never (n=36), transient (n=35), or persistent (n=14). After adjusting for height, children with persistent wheezing, compared to those who never wheezed, had -0.669 hPa·s·L ^-1^ XeE (95% confidence interval [CI] -1.102 to -0.237, p<0.01), -0.465 hPa·s·L ^-1^ ΔX (95%CI -0.772 to -0.159, p<0.01) and +1.433 hPa·s·L ^-1^ ΔX/V_T_ (95%CI +0.492 to +2.374, p<0.01). Increasing subject height had a significant effect on all IB-OSC resistance and reactance variables when adjusted for the effect of preschool wheezing.

**Conclusions:** IB-OSC is feasible for tracking lung function in preschool-aged children, and intra-breath reactance outcomes may allow abnormal lung function to be identified early in asymptomatic children with a history of persistent wheeze.

## Introduction

Longitudinal cohort studies have identified lower lung function, airway obstruction in early life, and poor lung function growth as risk factors for developing persistent asthma ^1-3^ and later chronic obstructive pulmonary disease. ^4,5^ Spirometry and plethysmography in healthy school-aged children have shown lung function increases in an ordered manner correlating with linear growth. ^6,7^ Epidemiological trajectory modelling techniques have identified several childhood risk factors for a low trajectory of the forced expiratory volume in 1-second (FEV _1_), including early wheezing, asthma, atopy, early childhood lower respiratory tract infections, and tobacco smoke exposure.^8,9^

Longitudinal studies of lung function in children under 6-years old relying on invoking expiratory flow limitation (EFL) using the rapid thoracoabdominal compression (RTC) technique or forced spirometry have several limitations. ^10-13^ First, the reliability and reproducibility of forced spirometry measurements in preschool-aged children are highly variable.^14^ Second, the lungs of healthy children under 6-years old are essentially empty within 1-second during a forced expiration manoeuvre, questioning the validity of FEV_1_ as an outcome variable in this age group. ^15^ Third, small airway disease is poorly reflected by spirometry.^16^ Fourth, infants with low maximal flow at functional residual capacity (V’ _maxFRC_) using the RTC technique have been found to have average or above-average FEV_1_ trajectories.^17^ It would be ideal to measure lung function trajectories in young children using an identical technique rather than comparing V’ _maxFRC_ and FEV _1_ to confirm the hypothesis that this is secondary to catch-up lung function growth.

Respiratory oscillometry is a non-invasive lung function technique that measures the impedance of the respiratory system (Zrs) during tidal breathing by superimposing periodic variations to transrespiratory pressure and measuring the response in oscillatory gas flow. ^18^ Within Zrs is biomechanical information about the resistance (Rrs) and reactance (Xrs) of the respiratory system where Rrs reflects airway and tissue resistive properties and Xrs reflects the elastic and inertive properties. ^18^ Oscillometry is highly feasible in preschoolers. ^19^ Previous oscillometry techniques – such as those using pseudorandom noise oscillations (spectral oscillometry) or recurrent impulses (impulse oscillometry) – average Zrs measurements across the entire respiratory cycle and are better suited to show between-group differences than to detect abnormal lung function in individual children. ^20,21^ Intra-breath oscillometry (IB-OSC) is a recent variant of oscillometry using a single-frequency sinusoidal oscillation to estimate Rrs and Xrs at discrete time points during the breathing cycle. By measuring Rrs and Xrs when the gas flow (V’) is zero (e.g., at moments of end-expiration and end-inspiration), the involvement of upper airway nonlinearities in Zrs can be minimised.^22^ The volume dependence of Rrs (ΔR) is calculated by measuring the difference between Rrs at end-expiration (ReE) and end-inspiration (ReI) and was shown by our group to detect airway obstruction in acutely wheezy children with a sensitivity of 92% and specificity of 89%.^21^

We aimed to assess the longitudinal changes in intra-breath Zrs in a cohort of clinically stable (i.e., asymptomatic) preschool-aged children. We hypothesised that the lung function growth trajectory in preschoolers with a history of wheezing would differ from that of healthy preschoolers who had never wheezed. We also hypothesised that the lung function growth trajectory in preschoolers with transient wheeze would be more like that of healthy preschoolers than those with persistent wheeze.

## Methods

### Study Subjects

This study was an extension of the Observational Research in Childhood Infectious Diseases (ORChID) community-based birth cohort study. ^23^ Children who participated in ORChID were invited for follow-up in the Early Life Lung Function (ELLF) study, which involved annual visits from ages 3 to 7 years. Demographic characteristics, height, weight, medical history, family history, respiratory symptoms, asthma risk factors, and environmental exposures were collected at each visit. Exclusion criteria for the ORChID study applicable to the ELLF study included chronic pulmonary (except asthma) or cardiovascular disease, chronic metabolic disorder, immunodeficiency, or children taking immunosuppressive medications. The Children’s Health Queensland (HREC/13/QRCH/156) and The University of Queensland (2013001291) Ethics Committees approved the study. Parents gave written, informed consent.

### Lung Function

IB-OSC was performed using a wave-tube oscillometry assembly designed and custom-made at the University of Szeged, Hungary, for “The International Collaboration to Improve Respiratory Health in Children” (INCIRCLE) European Respiratory Society (ERS) Clinical Research Collaboration ^24^ or the tremoflo® C-100, according to ERS technical standards. ^25^ The wave-tube methodology has been described previously. ^26^ The Zrs values are comparable for identical mechanical test loads between the INCIRCLE assembly and the tremoflo® C-100.^27^

A single 10 Hz sinusoid was used in 16–20 second measurements to track intra-breath changes in Zrs during tidal breathing. Test occasions containing at least three technically acceptable trials with a minimum of five breathing cycles containing no evidence of signal artefact from movement, coughing, vocalisation, glottic closure (e.g., swallowing), breath-holding, or leak from mouthpiece were included. All included recordings were analysed using the software of the custom-made oscillometer. Zrs variables determined by IB-OSC included resistance at end-expiration (ReE), resistance at end-inspiration (ReI), the tidal change in resistance (ΔR=ReE-ReI), reactance at end-expiration (XeE), reactance at end-inspiration (XeI), the tidal change in reactance (ΔX=XeE-XeI), and ΔX normalised by tidal volume (ΔX/V _T_). Individual breaths from each measurement trial were analysed, and variables were reported as the average of all technically acceptable measurements.

### Atopy

Atopy was defined by a positive skin prick test (≥3 mm wheal and greater than the positive histamine control) to at least one aeroallergen including cat, dog, house dust mite (*Dermatophagoides pteronyssinus* or *Dermatophagoides farinae*), mixed grasses, Bahia grass, *Alternaria tenuis*, and *Aspergillus fumigatus* performed at the final visit at 7-years of age.

### Wheeze groups

Wheeze groups were defined based on the wheezing phenotypes described in the Tucson Children’s Respiratory Study. ^10^ Subjects were divided into four groups based on parental reports. At each visit, parents were asked, *“Has your child ever had wheezing or whistling in the chest”* and *“Has your child had wheezing or whistling in the chest in the past 12 months?”* Based on responses, subjects were placed in one of four wheeze groups: never wheezed, transient wheeze, late-onset wheeze, or persistent wheeze (Table 1). Only one child met the definition of late-onset wheeze and was combined into the transient wheeze group to form three wheeze groups for analysis.

**Table 1.**
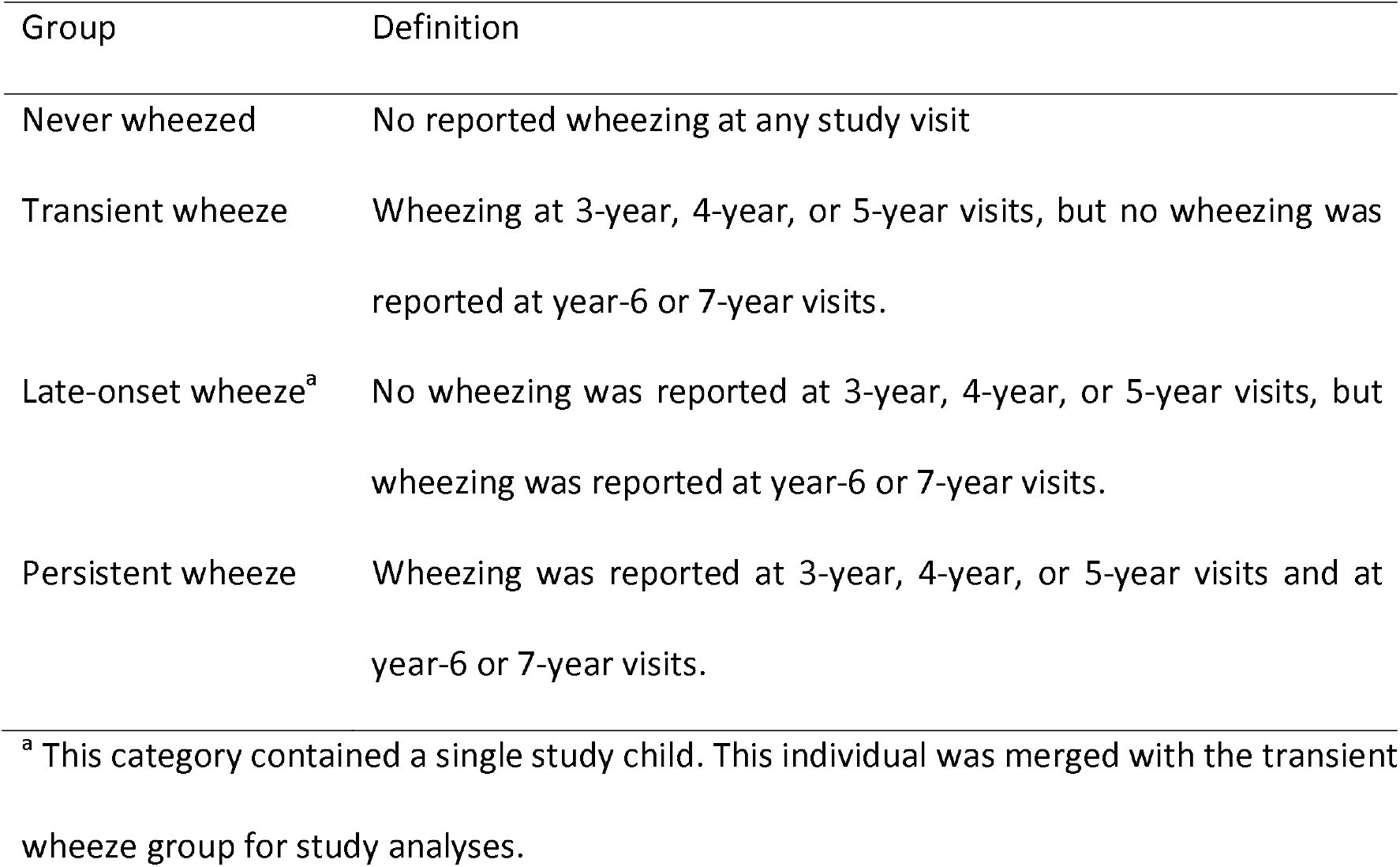
Wheeze groups based on the Tucson Children’s Respiratory Study.

| Group | Definition |
| --- | --- |
| Never wheezed | No reported wheezing at any study visit |
| Transient wheeze | Wheezing at 3-year, 4-year, or 5-year visits, but no wheezing was reported at year-6 or 7-year visits. |
| Late-onset wheeze<sup>a</sup> | No wheezing was reported at 3-year, 4-year, or 5-year visits, but wheezing was reported at year-6 or 7-year visits. |
| Persistent wheeze | Wheezing was reported at 3-year, 4-year, or 5-year visits and at year-6 or 7-year visits. |

<sup>a</sup> This category contained a single study child. This individual was merged with the transient wheeze group for study analyses.

### Statistical Analyses

Data are summarised as median (quartile-1, quartile-3) for continuous variables and as frequency and proportions for categorical variables. The coefficients of variability (CV) for ReE, ReI, XeE, and XeI were calculated for each visit according to the wheeze group. Normality was checked by conducting the Shapiro-Wilks test and visual inspection of histograms.

The association between wheeze groups and differences in IB-OSC outcomes over time was investigated using linear mixed-effects models (LMM). In all models, subjects were included as random effects to model between-subject variation. First, a null model was built for each IB-OSC outcome with no fixed effects. Then separate LMMs were fitted for each IB-OSC outcome with the wheeze group and height as fixed effects. LMMs were also fitted for each IB-OSC outcome with sex and height as fixed effects for the entire cohort. A sensitivity analysis was performed separately for subjects who never wheezed. We compared interaction models (height-by-wheeze group and height-by-sex) and non-interaction models for each outcome. We selected the most parsimonious model using the Akaike Information Criterion (AIC) (i.e., smallest AIC).^28^ Where appropriate, post-hoc analysis was performed for pairwise comparison between visits and wheeze groups using Bonferroni correction. All analyses were conducted using R statistical software (R Core Team, 2022) and the lmerTest package.^29^

## Results

Eighty-five children (54.1% female) attended visits across a 5-year follow-up period (Table 2). Thirty-six (42.4%) had never wheezed, 34 (40%) had transient wheeze (which included the single child with late-onset wheeze), and 14 (16.5%) had a history of persistent wheeze. The feasibility of IB-OSC measurements was 79.7% at 3-years, 93.8% at 4-years, 98.8% at 5-years, and 100% at 6 and 7-years (Table S1). Three hundred seventy-five test occasions (94.5% of attempted tests) yielded acceptable IB-OSC data (290 using the wave-tube assembly and 85 using the tremoflo® C-100) summarised in Table S2. The median coefficient of variability across all visits ranged from 12.4% to 13.5% for ReE, 12.9% to 13.9% for ReI, 50.9% to 68.2% for XeE, and 37.2% to 44.2% for XeI (Table S3).

**Table 2.** Early Life Lung Function (ELLF) study population characteristics.

| Visit age | 3-years | 4-years | 5-years | 6-years | 7-years |
| --- | --- | --- | --- | --- | --- |
|  | (n=82) | (n=84) | (n=85) | (n=81) | (n=81) |
| Females | 45 (54.9%) | 45 (53.6%) | 46 (54.1%) | 44 (54.3%) | 43 (53.1%) |
| n (%) |  |  |  |  |  |
| Height (cm)<sup>a</sup> | 98.1 | 104.4 | 111.1 | 117.9 | 124.2 |
|  | (96.0,100.6) | (102.7,107.0) | (109.0,114.1) | (115.1,120.4) | (120.9,127.8) |
| Wheezing |  |  |  |  |  |
| Never | 34 (41.5%) | 36 (42.9%) | 36 (42.4%) | 32 (39.5%) | 34 (42.0%) |
| Transient | 34 (41.5%) | 35 (41.7%) | 35 (41.2%) | 35 (43.2%) | 33 (40.7%) |
| Persistent | 14 (17.1%) | 13 (15.5%) | 14 (16.5%) | 14 (17.3%) | 14 (17.3%) |
| Household | 14 (17.1%) | 11 (13.1%) | 14 (16.5%) | 10 (12.3%) | 10 (12.3%) |
| Smoke |  |  |  |  |  |
| Exposure |  |  |  |  |  |
| Serum | 0.28 | 0.17 | n/a | n/a | 0.34 |
| eosinophils | (0.16,0.43) | (0.14,0.41) |  |  | (0.23,0.59) |
| (x10<sup>9</sup>/L)<sup>a</sup> |  |  |  |  |  |
| Atopy<sup>b</sup> | n/a | n/a | n/a | n/a | 24 (29.6%) |

Abbreviations: cm, centimetre; n/a, not applicable.
<sup>a</sup> Median (quartile-1, quartile-3).
<sup>b</sup> Based on skin prick testing at the 7-year of age visit.

The changes in intra-breath Zrs with growth (Figures 1 and 2) were explored using models containing: (1) subjects as random-effect and two predictors, height and wheeze group, as fixed-effects and (2) subjects as random-effect and height and sex as fixed-effects are presented below.

**Figure 1.**
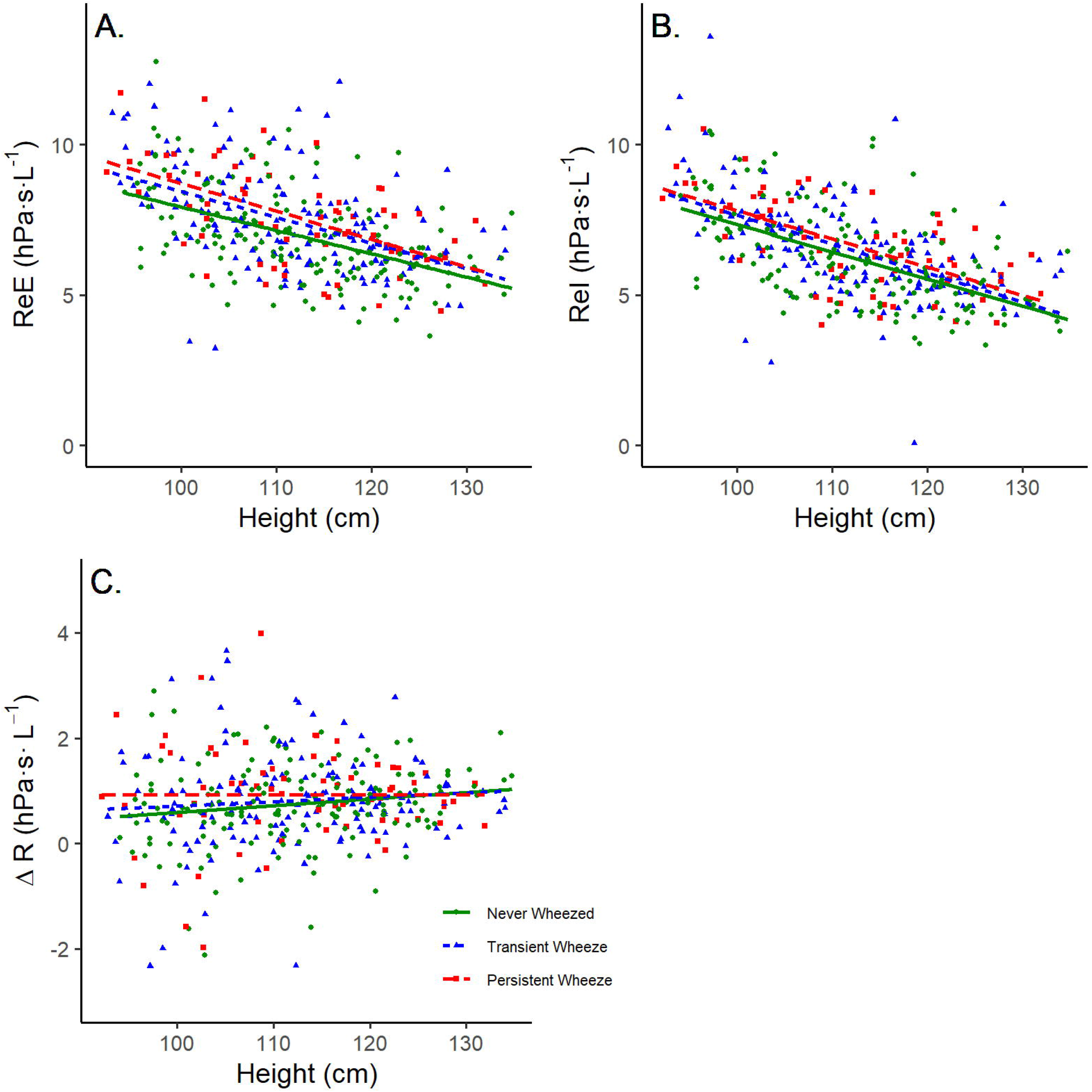
Longitudinal changes in (A) the resistance at end-expiration (ReE), (B) the resistance at end-inspiration (ReI), and (C) the tidal change in resistance (ΔR) with height.

**Figure 2.**
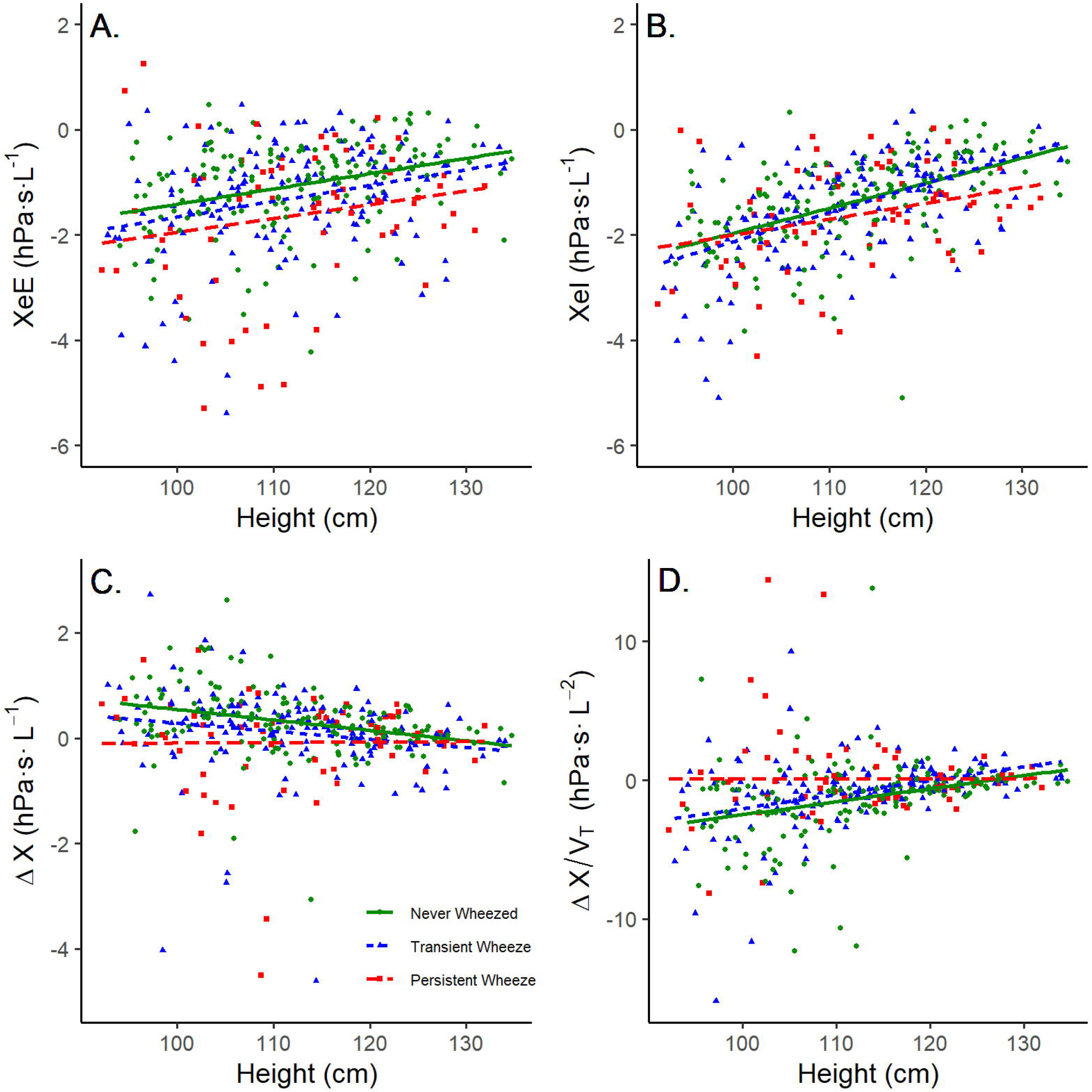
Longitudinal changes in (A) the reactance at end-expiration (XeE), (B) the reactance at end-inspiration (XeI), (C) the tidal change in reactance (ΔX), and (D) the tidal change in reactance normalised by tidal volume (ΔX/V_T_) with height.

### Association between subjects and longitudinal intra-breath Zrs

The results of the null models for each IB-OSC outcome are summarised in Table S4. Our final models with the height and wheeze group as fixed-effects indicate that in the height and wheeze group, 46.0% of the variance in ReE (intraclass correlation coefficient [ICC]=0.460), 30.8% of the variance in ReI (ICC=0.308), and 14.6% of the variance in ΔR (ICC=0.146) is explained by between-subject differences (Table S5). The ICC for reactance outcomes indicates that 28.6% of the variance in XeE, 15.5% of the variance in XeI, 4.2% of the variance in ΔX, and 11.0% of the variance in ΔX/V _T_ is explained by inter-subject differences.

### Association between linear growth and longitudinal intra-breath Zrs

The decrease in ReE, ReI, and ΔR per centimetre increase in height was -0.088 hectopascal per second per litre (hPa·s·L ^-1^) (95% confidence interval [CI] -0.100 to -0.076 p<0.001), - 0.098 hPa·s·L ^-1^ (95%CI -0.110 to -0.086, p<0.001), and +0.010 hPa·s·L ^-1^ (95%CI +0.001 to +0.019, p=0.035), respectively when the effect of wheezing was kept constant (Figure 1). The variation in outcome between subjects for ReE, ReI and ΔR was 0.96 standard deviations (SD), 0.74 SD, and 0.35 SD, respectively.

The changes in reactance for each centimetre increase in height were +0.034 hPa·s·L ^-1^ for XeE (95%CI +0.024 to +0.044, p<0.001), +0.050 hPa·s·L ^-1^ for XeI (95%CI +0.041 to +0.059, p<0.001), -0.011 hPa·s·L ^-1^ for ΔX (95%CI -0.021 to 0.000, p=0.046), and +0.078 hPa·s·L ^-2^ for ΔX/V_T_ (95%CI +0.050 to +0.107, p<0.001) when the effect of wheezing was kept constant (Figure 2). The variation in outcome between subjects for XeE, XeI, ΔX, and ΔX/V _T_ was 0.57 SD, 0.34 SD, 0.21 SD, and 0.93 SD, respectively.

### Association between wheezing and longitudinal intra-breath Zrs

After adjusting for height, the effect of wheezing on longitudinal intra-breath tracking of Zrs was statistically significant for XeE, ΔX, and ΔX/V _T_ but not XeI (Table 3). The subjects with transient wheeze, compared with subjects who never wheezed, had significantly worse (more negative) ΔX (mean difference [MD] -0.329, 95%CI -0.569 to -0.089, p=0.009). Subjects with persistent wheeze, compared with subjects who never wheezed, had significantly worse (more negative) XeE (MD -0.669, 95%CI -1.102 to -0.237, p=0.004) and ΔX (MD -0.465, 95%CI -0.772 to -0.159, p=0.004). The longitudinal change of ΔX/V _T_ was worse (more positive) by +1.433 hPa·s·L^-2^ (95%CI +0.492 to +2.374, p=0.004) in subjects with persistent wheeze than those who never wheezed. The wheezing phenotype had no significant effect on the longitudinal changes of ReE, ReI and ΔR after adjusting for height.

**Table 3.** Association of height and wheezing history with intra-breath impedance (n=375 observations from 85 children).

|  | Fixed effects |  | Estimate [mean (95%CI)] | p-value |
| --- | --- | --- | --- | --- |
| ReE<sup>†</sup> | Height (cm) |  | -0.088 (-0.100 to -0.076) | <0.001* |
|  | Wheeze Group<sup>§</sup> | Transient | +0.414 (-0.096 to +0.924) | 0.118 |
|  |  | Persistent | +0.586 (-0.075 to +1.258) | 0.088 |
| Rel<sup>†</sup> | Height (cm) |  | -0.098 (-0.110 to -0.086) | <0.001* |
|  | Wheeze Group<sup>§</sup> | Transient | +0.249 (-0.179 to +0.676) | 0.261 |
|  |  | Persistent | +0.412 (-0.140 to +0.965) | 0.151 |
| ΔR<sup>†</sup> | Height (cm) |  | +0.010 (+0.001 to +0.019) | 0.035* |
|  | Wheeze Group<sup>§</sup> | Transient | +0.109 (-0.145 to +0.363) | 0.405 |
|  |  | Persistent | +0.179 (-0.147 to +0.506) | 0.288 |
| XeE<sup>†</sup> | Height (cm) |  | +0.034 (+0.024 to +0.044) | <0.001* |
|  | Wheeze Group<sup>§</sup> | Transient | -0.245 (-0.580 to +0.090) | 0.159 |
|  |  | Persistent | -0.669 (-1.102 to -0.237) | 0.004* |
| Xel<sup>†</sup> | Height (cm) |  | +0.050 (+0.041 to +0.059) | <0.001* |
|  | Wheeze Group<sup>§</sup> | Transient | -0.006 (-0.248 to +0.236) | 0.962 |
|  |  | Persistent | -0.222 (-0.534 to +0.090) | 0.170 |
| $\Delta X^\dagger$ | Height (cm) | | -0.011 (-0.021 to 0.000) | 0.046<sup>*</sup> |
|  | Wheeze Group<sup>§</sup> | Transient | -0.329 (-0.569 to -0.089) | 0.009<sup>*</sup> |
|  |  | Persistent | -0.465 (-0.772 to -0.159) | 0.004<sup>*</sup> |
| $\Delta X/V_T^\ddagger$ | Height (cm) | | +0.078 (+0.050 to +0.107) | <0.001<sup>*</sup> |
|  | Wheeze Group<sup>§</sup> | Transient | +0.484 (-0.250 to +1.216) | 0.203 |
|  |  | Persistent | +1.433 (+0.492 to +2.374) | 0.004<sup>*</sup> |

Abbreviations: ReE, resistance at the end-expiration; Rel, resistance at end-inspiration; $\Delta R$ , tidal change in resistance ( $\Delta R = \text{ReE} - \text{Rel}$ ); XeE, reactance at end-expiration; Xel, reactance at end-inspiration; $\Delta X$ , tidal change in reactance ( $\Delta X = \text{XeE} - \text{Xel}$ ); $\Delta X/V_T$ , $\Delta X$ normalised by tidal volume.
<sup>\*</sup> $p < 0.05$
<sup>†</sup> Hectopascal per second per litre
<sup>‡</sup> Hectopascal per second per litre squared
<sup>§</sup> Reference category is the never wheezed group

### Association between sex and longitudinal intra-breath Zrs

When adjusting for the effect of height, there was no statistically significant effect of sex on the longitudinal changes of intra-breath Zrs or tidal volumes for the entire cohort (Table S6 and S7) or for subjects who never wheezed (Table S8).

## Discussion

Increasing height between the ages of 3-7 caused a reduction in ReE and ReI values and a decrease in the magnitude (less negative) of XeE, XeI and ΔX/V _T_ values. With linear growth, there were small but significant changes in both ΔR (more positive) and ΔX (more negative). When adjusting for the effect of height, persistent wheezers demonstrated a worse trajectory (more negative over time) of XeE, ΔX, and ΔX/V _T_ than subjects who never wheezed. The trajectory of ΔX across the preschool years was also worse in children with transient wheeze.

Our results are consistent with the height dependence of ReE, ReI, ΔR, XeE, XeI, and ΔX in healthy African children reported by Chaya et al. ^30^ Rrs represents the pressure changes in-phase with gas flow (V’) and reflects the sum of resistance from lung parenchyma and larger airways, where gas molecules move by bulk flow.^22^ In larger airways, Rrs due to laminar flow is inversely related to the radius of the airway to the fourth power (Poiseuille law). ^31^ Our observed trend of decreasing ReE and ReI with growth was expected as airway calibre increases by 200–300% from birth to adulthood. ^32^ Although ReE and ReI follow a similarly negative trajectory, these trajectories are not parallel and result in a small but significant increase in ΔR with preschool growth (Figure 1). The more gradual trajectory of ReE with height in this age group may reflect peripheral airway resistance making a greater contribution to the overall Rrs until the age of 5-years when conductance increases more rapidly.^33,34^

The observed trend of increasing (less negative) XeE and XeI with height (Figure 2) was also expected, as increasing lung volumes with growth would reduce the respiratory system elastance, which dominates Xrs at frequencies below the resonant frequency. The small but significant reduction in ΔX preschool-aged children may be due to the dysanaptic growth of the conducting airways compared to lung volumes up to 7-years of age.^34^ Airway calibre and length increase by a factor of 2-3 throughout childhood, while lung volume increases rapidly by a factor of 13 between birth and age 7-years. ^32,34^ Additionally, both ΔR and ΔX represent the volume dependence of Rrs and Xrs and may change in magnitude with larger tidal volumes in taller, healthy children (Table S8).

Czovek and colleagues found ΔR to be highly sensitive and specific for acute airway obstruction in preschool-aged children and distinguished those with recurrent wheezing from healthy children.^21^ In our data, ΔR was increased in transient and persistent wheezers, but changes were not statistically significant (Table 3). This may indicate that, with only 14 persistent wheezers, the study was underpowered to detect a true difference. The measurements in the present study were also made when children were asymptomatic. ΔR in asymptomatic children with a history of wheezing may not differ significantly from healthy children due to sparing of the larger calibre airways at an early age.

The longitudinal trajectory of ΔX in asymptomatic preschool children was significantly more negative in those with a history of wheezing when the effect of growth was kept constant (Figure 2). Changes in Xrs reflect elastic properties of the lung and thus are influenced by the ventilation of individual lung units. Time-constant inhomogeneity, produced by changes in the mechanical properties of the smaller airways, will induce a shift in XeE to become more negative as some units are under ventilated while others are over ventilated, increasing overall lung stiffness. ^18,35^ The abnormal (more negative) trajectory of ΔX in our cohort of children with transient and persistent wheeze may reflect an evolution of ventilation inhomogeneity over time in wheezy children that does not improve with growth in lung volumes.^21^

In a cross-sectional study of adults with asthma, baseline impedance data including ReE, ReI, ΔR, XeE, XeI, and ΔX distinguished those with controlled versus uncontrolled asthma. ^36^ However, spirometry variables were unable to distinguish the difference between these two groups. In our cohort of preschool-aged children, an abnormal trajectory for ΔX may be the earliest sign of small airway dysfunction in the lung periphery.^37^

We report a novel outcome ΔX/V _T_, which normalises the change in ΔX for tidal volume. Healthy children with no airway disease tend to have no change or a slight decrease in Xrs between end expiration and end inspiration (ΔX≥0). In small airway disease, the higher elastance at end-expiration (more negative XeE values) indicates mechanical inhomogeneity of peripheral lung units, which improves with inspiration (increasing Xrs) and manifests in negative ΔX and ΔX/V_T_ values.^21,35^ Our longitudinal data support this theory by showing that the trajectory of ΔX/V _T_ over time in preschool-aged children with persistent wheeze becomes significantly more positive than in those who have never wheezed (Table 3). This new variable may have clinical significance in other diseases, such as cystic fibrosis, where measurements of ventilation inhomogeneity are important for identifying early lung disease progression but often challenging to achieve in a clinical setting for young children.^38^

Our study has several strengths. This is the first longitudinal study using IB-OSC in a community-based cohort of clinically stable preschool-aged children. IB-OSC was highly feasible with acceptable longitudinal Zrs data obtained from 79.7% of lung function naïve children aged 3-years and 98–100% of subjects 5-years and older. This is similar to the high longitudinal feasibility of impulse oscillometry in children aged 4 to 6-years reported previously^39^ and significantly higher than the longitudinal feasibility of impulse oscillometry (21% at 4-years, 58% at 5-years, 74% at 6-years, 79% at 7-years, 86% at 8-years) which may reflect differences in tolerance of a single sinusoid compared to an impulse waveform.^14^ Our feasibility is comparable to cross-sectional studies reporting the feasibility of oscillometry in preschool-aged children.^40^

We have demonstrated that IB-OSC can track lung function trajectories using identical methods and outcomes from age 3-years onwards instead of relying on different paediatric outcomes like maximum flow at functional residual capacity (V’maxFRC) or forced expiratory volume in 0.5 s (FEV_0.5_) which are not useful parameters in older children and adults.

All subjects were part of a birth cohort study with non-random recruitment, which may contribute to selection bias, but does not change the internal validity of our study results. Our wheeze groups were based on parental annual reports of wheezing in the previous 12 months, which may be subject to responder error. Although we used mixed-effects models to account for subject and growth effects, our sample size was too small for more complex multivariable models.

This study confirms that IB-OSC is feasible for tracking longitudinal lung function in preschool-aged children. Asymptomatic young children with a history of persistent wheeze have significantly altered lung function trajectories for XeE, ΔX, and ΔX/V _T_ compared to children who have never wheezed after adjusting for the effect of linear growth. Our study suggests that intra-breath reactance outcomes may be helpful for the early identification of children with abnormal lung function in a clinical setting.

## Author Contributions

Matthew D. Wong: Conceptualization (supporting); data curation (lead); formal analysis (lead); methodology (lead); validation (lead); visualization (lead); writing – original draft (lead); writing – review & editing (lead).

Tamara Blake: Conceptualization (supporting); data curation (supporting); investigation (supporting); project administration (supporting); resources (supporting); supervision (equal); writing – original draft (supporting); writing – review & editing (supporting).

Syeda Farah Zahir: Conceptualization (supporting); data curation (supporting); formal analysis (supporting); methodology (supporting); supervision (equal); validation (supporting); visualization (supporting); writing – original draft (supporting); writing – review & editing (supporting).

Sadasivam Suresh: Conceptualization (supporting); formal analysis (supporting); supervision (equal); writing – original draft (supporting); writing – review & editing (supporting).

Zoltán Hantos: Conceptualization (supporting); formal analysis (supporting); funding acquisition (supportive); resources (supporting); supervision (equal); validation (supporting); writing – original draft (supporting); writing – review & editing (supporting).

Keith Grimwood: Conceptualization (lead); funding acquisition (equal); validation (supporting); writing – review & editing (supporting).

Stephen B. Lambert: Conceptualization (lead); funding acquisition (equal); validation (supporting); writing – review & editing (supporting).

Robert S. Ware: Conceptualization (lead); funding acquisition (equal); validation (supporting); writing – review & editing (supporting).

Peter D. Sly: Conceptualization (lead); data curation (supporting); formal analysis (supporting); funding acquisition (equal); investigation (supporting); methodology (supporting); project administration (supporting); resources (lead); supervision (lead); validation (supporting); writing – original draft (supporting); writing – review & editing (supporting).

## Data Availability

The data that support the findings of this study are available on request from the corresponding author, MDW. The data are not publicly available due to privacy/ethical reasons.

## Acknowledgements

We thank Dr Mohammad Zahirul Islam and Ms Sally Galbraith for their support of the study and Prof Paul Robinson for his editorial suggestions. The authors also thank the families and children who participated in the study.

## Research funding

This study was supported by the National Health and Medical Research Council (GNT1078660). ZH was supported by Hungarian Scientific Research Fund (Grant K 128701). ZH and PDS were supported by the European Respiratory Society Clinical Research Collaboration Award (CRC_2013-02_INCIRCLE).

## Human ethics approval declaration

This study was performed in accordance with the Declaration of Helsinki and approved by the Children’s Health Queensland (HREC/13/QRCH/156) and The University of Queensland (2013001291) Ethics Committees. All parents, guardians or next of kin provided written informed consent for all study subjects.

