## Supplementary material for "Longitudinal tracking of intra-breath respiratory impedance in preschool children": Table S1

Table S1. Feasibility of intra-breath oscillometry (IB-OSC) outcomes by age

| IB-OSC | Total | 3-years | 4-years | 5-years | 6-years | 7-years |
|---|---|---|---|---|---|---|
| Attempted | 397 | 79 | 80 | 82 | 78 | 78 |
| Acceptable | 375 | 63 | 75 | 81 | 78 | 78 |
| Feasibility* | | 79.7% | 93.8% | 98.8% | 100% | 100% |

Note: Feasibility = $\left(\frac{number\ of\ children\ with\ acceptable\ IB{-}OSC\ testing}{number\ of\ children\ who\ attempted\ IB{-}OSC\ testing}\right) \times 100\%$
