## Supplementary material for "Longitudinal tracking of intra-breath respiratory impedance in preschool children": Table S2

Table S2. Summary of IB-OSC Zrs data based on wheeze phenotype and visit.

| | Wheeze group | 3-year visit* | 4-year visit* | 5-year visit* | 6-year visit* | 7-year visit* |
|---|---|---|---|---|---|---|
| ReE† | *Never* | +8.48 [+7.18, +9.56] | +7.22 [+6.69, +8.69] | +6.75 [+6.01, +7.50] | +6.29 [+5.67, +7.08] | +5.93 [+5.44, +6.31] |
| | *Transient* | +8.81 [+8.20, +9.95] | +7.78 [+7.00, +8.92] | +7.18 [+6.33, +8.06] | +6.70 [+5.78, +7.35] | +6.41 [+6.03, +6.97] |
| | *Persistent* | +9.26 [+8.49, +9.65] | +7.94 [+7.09, +9.02] | +6.64 [+5.70, +7.90] | +7.28 [+6.39, +7.71] | +6.73 [+6.18, +7.64] |
| ReI† | *Never* | +7.63 [+6.94, +8.61] | +6.82 [+5.91, +8.27] | +6.14 [+5.11, +6.94] | +5.47 [+4.75, +6.24] | +4.84 [+4.73, +5.49] |
| | *Transient* | +8.14 [+7.50, +9.28] | +6.89 [+6.44, +7.66] | +6.19 [+5.68, +7.22] | +5.56 [+5.24, +6.37] | +5.56 [+5.08, +6.18] |
| | *Persistent* | +8.64 [+8.10, +8.76] | +7.60 [+6.48, +8.14] | +5.98 [+4.74, +6.97] | +5.74 [+5.18, +6.33] | +6.07 [+5.14, +6.68] |
| ΔR† | *Never* | +0.65 [+0.02, +1.10] | +0.56 [+0.13, +0.95] | +0.79 [+0.22, +1.46] | +0.72 [+0.54, +1.10] | +1.00 [+0.76, +1.29] |
| | *Transient* | +0.55 [+0.35, +1.02] | +0.82 [+0.25, +1.72] | +0.64 [+0.21, +1.25] | +0.76 [+0.38, +1.24] | +0.86 [+0.64, +1.12] |
| | *Persistent* | +0.73 [-0.23, +1.90] | +1.06 [+0.31, +1.42] | +0.98 [+0.72, +1.20] | +1.21 [+0.90, +1.56] | +0.79 [+0.40, +1.12] |
| XeE† | *Never* | -1.45 [-1.98, -0.87] | -1.12 [-1.59, -0.62] | -0.97 [-1.62, -0.40] | -0.73 [-0.92, -0.48] | -0.56 [-0.86, -0.19] |
| | *Transient* | -2.02 [-2.71, -1.62] | -1.22 [-1.93, -0.94] | -0.97 [-1.59, -0.49] | -0.84 [-1.43, -0.48] | -0.93 [-1.51, -0.40] |
| | *Persistent* | -1.83 [-2.67, -0.92] | -2.09 [-3.73, -0.96] | -0.95 [-1.43, -0.62] | -1.40 [-1.92, -0.92] | -1.19 [-1.84, -0.82] |
| XeI† | *Never* | -2.02 [-2.45, -1.58] | -1.79 [-2.24, -1.25] | -1.40 [-1.93, -0.85] | -0.94 [-1.16, -0.68] | -0.61 [-1.08, -0.26] |
| | *Transient* | -2.53 [-3.37, -1.84] | -1.69 [-1.95, -1.36] | -1.10 [-1.88, -0.78] | -0.87 [-1.22, -0.66] | -0.88 [-1.27, -0.50] |

| | | | | | | |
|---|---|---|---|---|---|---|
| | *Persistent* | -2.17 [-2.73, -1.57] | -2.50 [-2.94, -1.38] | -1.32 [-1.87, -0.81] | -1.29 [-1.72, -0.67] | -1.31 [-1.48, -0.84] |
| ΔX† | *Never* | +0.49 [+0.13, +0.94] | +0.57 [+0.21, +0.86] | +0.32 [-0.09, +0.66] | +0.24 [-0.02, +0.42] | +0.08 [-0.08, +0.21] |
| | *Transient* | +0.28 [-0.10, +0.58] | +0.34 [-0.19, +0.62] | +0.29 [+0.05, +0.51] | +0.20 [+0.04, +0.42] | -0.12 [-0.32, +0.13] |
| | *Persistent* | +0.51 [-0.03, +0.79] | +0.07 [-0.69, +0.28] | +0.31 [+0.14, +0.50] | -0.04 [-0.55, +0.39] | +0.02 [-0.29, +0.17] |
| $\Delta X/V_T$‡ | *Never* | -2.23 [-3.51, -0.59] | -2.35 [-4.30, -0.85] | -0.96 [-2.17, +0.18] | -0.52 [-1.14, +0.06] | -0.15 [-0.47, +0.16] |
| | *Transient* | -1.00 [-3.39, -0.23] | -1.34 [-2.37, +0.42] | -0.85 [-1.80, -0.14] | -0.46 [-0.80, -0.05] | +0.24 [-0.32, +0.79] |
| | *Persistent* | -2.05 [-3.55, +0.14] | -0.09 [-1.14, +1.77] | -1.24 [-1.62, -0.41] | +0.12 [-0.67, +1.43] | -0.05 [-0.50, +0.73] |
| $V_T$ (L) | All subjects | +0.24 [+0.20, +0.30] | +0.28 [+0.19, +0.34] | +0.30 [+0.24, +0.40] | +0.41 [+0.34, +0.49] | +0.43 [+0.36, +0.52] |

Abbreviations: IB-OSC, intra-breath oscillometry; L, litres; ReE, resistance at end-expiration; ReI, resistance at end-inspiration; ΔR, tidal change in resistance (ΔR=ReE-ReI); XeE, reactance at end-expiration; XeI, reactance at end-inspiration; ΔX, tidal change in reactance (ΔX=XeE-XeI); $\Delta X/V_T$, ΔX normalised by tidal volume; $V_T$, tidal volume; Zrs, intra-breath respiratory impedance.

* Median [quartile-1,quartile-3]

† Hectopascal per second per litre

‡ Hectopascal per second per litre squared
