## Supplementary material for "Longitudinal tracking of intra-breath respiratory impedance in preschool children": Table S3

Table S3. Coefficient of variability for ReE, ReI, XeE, and XeI

| | Wheeze group | 3-year visit* | 4-year visit* | 5-year visit* | 6-year visit* | 7-year visit* | Total* |
|---|---|---|---|---|---|---|---|
| ReE | *Never* | 12.7 [9.4, 18.1] | 14.1 [10.7, 19.5] | 13.6 [10.8, 16.1] | 10.9 [9.2, 14.3] | 11.3 [9.6, 14.1] | 12.9 [9.8, 16.2] |
| | *Transient* | 13.4 [10.3, 19.8] | 14.2 [11.6, 18.4] | 14.3 [9.9, 18.4] | 12.1 [10.4, 16.4] | 12.4 [10.7, 15.1] | 13.5 [10.5, 17.7] |
| | *Persistent* | 14.1 [11.1, 18.3] | 14.6 [13.0, 19.7] | 10.7 [8.1, 14.2] | 11.5 [8.6, 17.4] | 11.5 [8.8, 13.6] | 12.4 [9.0, 16.3] |
| ReI | *Never* | 13.0 [10.2, 18.9] | 13.1 [10.5, 17.8] | 13.5 [10.0, 17.5] | 14.1 [10.7, 16.8] | 14.9 [11.7, 16.6] | 13.9 [10.8, 17.5] |
| | *Transient* | 14.2 [11.1, 20.2] | 15.6 [11.8, 21.2] | 11.8 [9.8, 15.6] | 12.8 [11.4, 16.8] | 13.4 [11.5, 16.1] | 13.3 [11.0, 18.3] |
| | *Persistent* | 13.4 [11.2, 17.4] | 13.3 [12.4, 15.7] | 10.4 [8.7, 13.4] | 15.1 [9.8, 18.5] | 12.0 [10.1, 14.8] | 12.9 [9.8, 15.7] |
| XeE | *Never* | 58.3 [52.0, 78.3] | 69.3 [55.9, 140.2] | 52.5 [42.8, 107.2] | 59.9 [43.7, 121.3] | 79.4 [51.9, 132.1] | 62.5 [48.2, 111.6] |
| | *Transient* | 52.8 [40.1, 84.2] | 65.0 [47.5, 122.9] | 67.6 [49.7, 126.0] | 75.3 [45.6, 135.1] | 75.2 [42.6, 133.1] | 68.2 [43.9, 119.3] |
| | *Persistent* | 42.6 [36.8, 80.3] | 53.3 [47.3, 105.7] | 55.4 [34.3, 72.0] | 48.0 [33.1, 77.1] | 54.7 [36.0, 81.2] | 50.9 [36.0, 84.0] |
| XeI | *Never* | 34.0 [24.4, 44.0] | 45.9 [34.9, 58.3] | 31.4 [25.3, 45.8] | 39.3 [30.0, 61.6] | 51.3 [37.6, 108.8] | 39.9 [28.4, 59.9] |
| | *Transient* | 36.2 [23.2, 51.4] | 41.3 [31.3, 53.0] | 38.7 [25.9, 66.7] | 44.8 [29.3, 62.6] | 62.1 [35.7, 94.9] | 44.2 [29.6, 65.4] |
| | *Persistent* | 50.2 [30.6, 62.4] | 35.8 [29.6, 52.5] | 40.5 [28.1, 45.9] | 31.4 [20.4, 90.8] | 34.7 [27.0, 63.6] | 37.2 [27.2, 58.9] |

Abbreviations: ReE, resistance at end-expiration; ReI, resistance at end-inspiration; XeE, reactance at end-expiration; XeI, reactance at end-inspiration.

* Median % [quartile-1, quartile-3]
