## Supplementary material for "Longitudinal tracking of intra-breath respiratory impedance in preschool children": Table S4

Table S4. Null linear mixed-effects models for each IB-OSC outcome using subjects as random-effects and no fixed-effects.

| | Estimated intercept | Estimated variance of random effect | Intraclass Correlation Coefficient (ICC) |
|---|---|---|---|
| ReE† | 7.238 | 0.847 | 0.312 |
| ReI† | 6.406 | 0.385 | 0.147 |
| ΔR† | 0.818 | 0.120 | 0.139 |
| XeE† | -1.252 | 0.326 | 0.260 |
| XeI† | -1.462 | 0.056 | 0.057 |
| ΔX† | 0.172 | 0.072 | 0.069 |
| $\Delta X/V_T$‡ | -0.875 | 1.079 | 0.125 |

Abbreviations: IB-OSC, intra-breath oscillometry; ReE, resistance at end-expiration; ReI, resistance at end-inspiration; ΔR, tidal change in resistance (ΔR=ReE-ReI); XeE, reactance at end-expiration; XeI, reactance at end-inspiration; ΔX, tidal change in reactance (ΔX=XeE-XeI); $\Delta X/V_T$, ΔX normalised by tidal volume.

† Hectopascal per second per litre

‡ Hectopascal per second per litre squared
