## Supplementary material for "Longitudinal tracking of intra-breath respiratory impedance in preschool children": Table S5

Table S5. Final linear mixed-effects models for each IB-OSC outcome using subjects as random-effects and fixed-effects of height and wheeze group.

| | Intraclass Correlation Coefficient (ICC) |
|---|---|
| ReE[†] | 0.460 |
| ReI[†] | 0.308 |
| ΔR[†] | 0.146 |
| XeE[†] | 0.286 |
| XeI[†] | 0.155 |
| ΔX[†] | 0.042 |
| $\Delta X/V_T$[‡] | 0.110 |

Abbreviations: IB-OSC, intra-breath oscillometry; ReE, resistance at end-expiration; ReI, resistance at end-inspiration; ΔR, tidal change in resistance (ΔR=ReE-ReI); XeE, reactance at end-expiration; XeI, reactance at end-inspiration; ΔX, tidal change in reactance (ΔX=XeE-XeI); $\Delta X/V_T$, ΔX normalised by tidal volume.

[†] Hectopascal per second per litre

[‡] Hectopascal per second per litre squared
