## Supplementary material for "Longitudinal tracking of intra-breath respiratory impedance in preschool children": Table S6

Table S6. Association of height and sex with longitudinal intra-breath Zrs.

| | Fixed effects | | Estimate [mean (95%CI)] | *p*-value |
|---|---|---|---|---|
| ReE† | Height (cm) | | -0.087 (-0.099 to -0.075) | <0.001* |
| | Sex | Female | +0.330 (-1.909 to +2.581) | 0.774 |
| | | Male | +0.016 (-2.259 to +2.303) | 0.989 |
| ReI† | Height (cm) | | -0.097 (-0.109 to -0.084) | <0.001* |
| | Sex | Female | +1.498 (-0.846 to +3.840) | 0.212 |
| | | Male | +1.169 (-1.193 to +3.529) | 0.333 |
| ΔR† | Height (cm) | | +0.010 (+0.001 to +0.019) | 0.038* |
| | Sex | Female | -0.803 (-2.561 to +0.986) | 0.373 |
| | | Male | -0.861 (-2.624 to +0.933) | 0.341 |
| XeE† | Height (cm) | | +0.034 (+0.024 to +0.044) | <0.001* |
| | Sex | Female | -0.333 (-2.256 to +1.566) | 0.732 |
| | | Male | -0.572 (-2.509 to +1.342) | 0.560 |
| XeI† | Height (cm) | | +0.050 (+0.041 to +0.058) | <0.001* |
| | Sex | Female | -0.468 (-2.145 to +1.193) | 0.583 |
| | | Male | -0.425 (-2.106 to +1.241) | 0.619 |

| | | | | |
|---|---|---|---|---|
| ΔX[†] | Height (cm) | | -0.009 (-0.020 to 0.001) | 0.088 |
| | Sex | Female | -0.203 (-2.202 to +1.771) | 0.841 |
| | | Male | -0.497 (-2.496 to +1.479) | 0.624 |
| $\Delta X/V_T$[‡] | Height (cm) | | +0.076 (+0.047 to +0.104) | <0.001* |
| | Sex | Female | -0.258 (-5.658 to +5.221) | 0.926 |
| | | Male | +0.482 (-4.931 to +5.974) | 0.862 |
| $V_T$ (L) | Height (cm) | | +0.012 (-0.003 to +0.026) | 0.114 |
| | Sex | Female | -0.072 (-2.837 to +2.690) | 0.959 |
| | | Male | +0.115 (-2.648 to +2.877) | 0.935 |

Abbreviations: CI, confidence interval; cm, centimetres; L, litres; ReE, resistance at end-expiration; ReI, resistance at end-inspiration; ΔR, tidal change in resistance (ΔR=ReE-ReI); XeE, reactance at end-expiration; XeI, reactance at end-inspiration; ΔX, tidal change in reactance (ΔX=XeE-XeI); $\Delta X/V_T$, ΔX normalised by tidal volume; $V_T$, tidal volume; Zrs, respiratory impedance.

* $p < 0.05$

† Hectopascal per second per litre

‡ Hectopascal per second per litre squared
