## Supplementary material for "Longitudinal tracking of intra-breath respiratory impedance in preschool children": Table S7

Table S7. Final linear mixed-effects models for each IB-OSC outcome using subjects as random-effects and fixed-effects of sex and height.

| | Adjusted Intraclass Correlation Coefficient |
|---|---|
| ReE[†] | 0.464 |
| ReI[†] | 0.304 |
| ΔR[†] | 0.152 |
| XeE[†] | 0.306 |
| XeI[†] | 0.156 |
| ΔX[†] | 0.052 |
| $\Delta X/V_T$[‡] | 0.119 |
| $V_T$ (L) | 0.007 |

Abbreviations: IB-OSC, intra-breath oscillometry; L, litres; ReE, resistance at end-expiration; ReI, resistance at end-inspiration; ΔR, tidal change in resistance (ΔR=ReE-ReI); XeE, reactance at end-expiration; XeI, reactance at end-inspiration; ΔX, tidal change in reactance (ΔX=XeE-XeI); $\Delta X/V_T$, ΔX normalised by tidal volume; $V_T$, tidal volume.

[†] Hectopascal per second per litre

[‡] Hectopascal per second per litre squared
