## Supplementary material for "Longitudinal tracking of intra-breath respiratory impedance in preschool children": Table S8

Table S8. Association of height and sex with longitudinal intra-breath Zrs for subjects who have never wheezed.

| | Fixed effects | | Estimate [mean (95%CI)] | *p*-value |
|---|---|---|---|---|
| ReE† | Height (cm) | | -0.083 (-0.100 to -0.066) | <0.001* |
| | Sex | Female | +0.347 (-1.784 to +2.502) | 0.752 |
| | | Male | -0.165 (-2.382 to +2.072) | 0.885 |
| ReI† | Height (cm) | | -0.098 (-0.114 to -0.081) | <0.001* |
| | Sex | Female | +1.477 (-0.595 to +3.542) | 0.165 |
| | | Male | +1.202 (-0.952 to +3.350) | 0.278 |
| ΔR† | Height (cm) | | +0.015 (+0.003 to +0.026) | 0.015* |
| | Sex | Female | -0.819 (-2.245 to +0.679) | 0.267 |
| | | Male | -1.059 (-2.503 to +0.449) | 0.156 |
| XeE† | Height (cm) | | +0.030 (+0.015 to +0.044) | <0.001* |
| | Sex | Female | -0.669 (-2.522 to +1.088) | 0.460 |
| | | Male | -0.648 (-2.502 to +1.121) | 0.478 |
| XeI† | Height (cm) | | +0.049 (+0.035 to +0.062) | <0.001* |
| | Sex | Female | -0.470 (-2.114 to +1.134) | 0.571 |
| | | Male | -0.348 (-2.001 to +1.272) | 0.678 |

| | | | | |
|---|---|---|---|---|
| $\Delta X^{\dagger}$ | Height (cm) | | -0.018 (-0.035 to -0.001) | 0.042* |
| | Sex | Female | -0.319 (-2.398 to +1.760) | 0.766 |
| | | Male | -0.427 (-2.508 to +1.654) | 0.690 |
| $\Delta X/V_T^{\ddagger}$ | Height (cm) | | +0.094 (+0.049 to +0.138) | <0.001* |
| | Sex | Female | +0.670 (-4.780 to +6.119) | 0.811 |
| | | Male | +0.807 (-4.646 to +6.261) | 0.774 |
| $V_T$ (L) | Height (cm) | | +0.009 (+0.006 to +0.011) | <0.001* |
| | Sex | Female | -0.102 (-0.356 to +0.153) | 0.438 |
| | | Male | -0.100 (-0.356 to +0.156) | 0.448 |

Abbreviations: CI, confidence interval; cm, centimetres; L, litres; ReE, resistance at end-expiration; ReI, resistance at end-inspiration; ΔR, tidal change in resistance (ΔR=ReE-ReI); XeE, reactance at end-expiration; XeI, reactance at end-inspiration; ΔX, tidal change in reactance (ΔX=XeE-XeI); $\Delta X/V_T$, ΔX normalised by tidal volume; $V_T$, tidal volume; Zrs, respiratory impedance.

* $p < 0.05$

† Hectopascal per second per litre

‡ Hectopascal per second per litre squared
